# MenstLLaMA: A Specialized Large Language Model for Menstrual Health Education in India

**DOI:** 10.1101/2025.01.22.25320466

**Authors:** Prottay Kumar Adhikary, Isha Motiyani, Gayatri Oke, Maithili Joshi, Kanupriya Pathak, Salam Michael Singh, Tanmoy Chakraborty

## Abstract

**Background:** The quality and accessibility of menstrual health education in developing nations, including India, remain inadequate due to challenges such as poverty, social stigma, and gender inequality. While community-driven initiatives aim to raise awareness, artificial intelligence (AI) offers a scalable solution for disseminating accurate information. However, existing general-purpose large language models (LLMs) are ill-suited for this task, suffering from low accuracy, cultural insensitivity, and overly complex responses. To address these limitations, we developed MenstLLaMA, a specialized LLM tailored to the Indian context, designed to deliver menstrual health education empathetically, supportively, and accessible.

**Objective:** To develop and evaluate MenstLLaMA, a specialized LLM tailored to deliver accurate, culturally sensitive menstrual health education, and to assess its effectiveness compared to existing general-purpose models.

**Methods:** We curated MENST, a novel domain-specific dataset comprising 23,820 question-answer pairs, aggregated from medical websites, government portals, and health education resources. This dataset was systematically annotated with metadata capturing age groups, regions, topics, and socio-cultural contexts. MenstLLaMA was developed by fine-tuning Meta-LLaMA-3-8B-Instruct using Parameter Efficient Fine-Tuning (PEFT) with Low-Rank Adaptation (LoRA) techniques to achieve domain alignment with reduced computational overhead. We benchmarked MenstLLaMA against nine state-of-the-art general-purpose LLMs, including GPT-4o, Claude-3, Gemini 1.5 Pro, Mistral, and others. The evaluation followed a multi-layered framework: (1) automatic evaluation using BLEU, METEOR, ROUGE-L, and BERTScore metrics (2) expert evaluation by clinical experts (N=18) rating 200 expert-curated queries (3) medical practitioner interaction using an interactive chatbot (ISHA) for qualitative assessment across *Relevance, Understandability, Preciseness, Correctness* and *Context sensitivity* and (4) a user study with volunteer participants (N=200) evaluating MenstLLaMA in 15–20 minute randomized sessions for user satisfaction assessment on performance across seven qualitative metrics.

**Results:** MenstLLaMA achieved the highest BLEU (0.059) and BERTScore (0.911), outperforming GPT-4o (BLEU: 0.052, BERTScore: 0.896) and Claude-3 (BERTScore: 0.888). Clinical experts preferred MenstLLaMA’s responses over gold-standard answers in several culturally sensitive cases. In evaluation by medical practitioners ISHA, the chat interface of MenstLLaMA, it scored 3.5 in Relevance, 3.6 in *Understandability*, 3.1/5 in *Preciseness*, 3.5/5 in *Correctness*, and 4.0/5 in *Context Sensitivity*. User evaluations indicated strong ratings for *Understandability* (4.7/5), *Relevance* (4.3/5), *Preciseness* (4.28/5), *Correctness* (4.1/5), *Tone* (4.6/5), *Flow* (4.2/5), and *Context Sensitivity* (3.9/5).

**Conclusions:** MenstLLaMA demonstrates exceptional accuracy, empathy, and user satisfaction in menstrual health education, bridging critical gaps left by general-purpose LLMs. Its potential for integration into broader health education platforms positions it as a transformative tool for menstrual well-being. Future research may explore its long-term impact on public perception, menstrual hygiene practices, expanding demographic representation, enhancing context sensitivity, and integrating multi-modal and voice-based interactions for broader accessibility.

## Introduction

### Overview

Menstruation, a biological process experienced by nearly half the global population, continues to be shrouded in stigma and misinformation, particularly in low-income and rural areas [1]. This accentuates the lack of awareness about menstrual hygiene, resulting in health risks and contributing to the spread of misinformation. In India, menstrual health remains a deeply ingrained taboo across both rural and urban settings [2]. Women are often segregated during menstruation, barred from entering places of worship, and labelled as “impure” or “unclean” during this natural cycle [3,4]. This societal ostracism and inadequate menstrual health education (MHE) have far-reaching consequences. For instance, nearly 24% of school-aged girls in India skip or drop out of school altogether due to menstruation-related issues [5], reflecting the persistent discrimination they face from a young age [2]. These challenges underscore the urgent need for nationwide initiatives to make MHE accessible, inclusive, and culturally sensitive.

MHE can empower women and benefit society in multiple ways. It can impart essential information about maintaining menstrual hygiene and accessing supplies and sanitation facilities to prevent menstrual health disorders [6]. Simultaneously, it can address the social stigma surrounding menstruation and help dismantle cultural taboos that restrict women from fully participating in various spheres of life while they are menstruating [6]. While India’s education system has begun incorporating MHE into school curricula, these efforts often fall short, failing to address cultural sensitivities or challenge the deeply rooted menstrual stigma in the society [7]. Non-governmental organizations (NGOs) have stepped in to bridge this gap, conducting community-based workshops and awareness campaigns nationwide [7]. However, the advent of Generative Artificial Intelligence (Gen AI) and Large Language Models (LLMs) offers an unprecedented opportunity to scale these efforts and reshape public understanding of menstrual health.

Most Indian population, particularly the youth, has internet access and relies on it as a primary knowledge source. However, while the internet offers a vast repository of information, it is also prone to spreading misinformation and adding to the stigma associated with menstruation [8]. The introduction of LLMs offer new possibilities for disseminating accurate menstrual health information and combating misinformation [9]. Compared to simpler interventions like static mobile apps or traditional chatbots, LLMs provide dynamic, conversational, and context-aware responses that can adapt to user queries more naturally and empathetically. This adaptability is particularly important in culturally sensitive topics like menstruation, where rigid or templated responses may fall short. Moreover, a single LLM-based solution can serve diverse linguistic and contextual needs without requiring extensive hard-coded rules, making it a scalable tool even in low-resource settings where specialized infrastructure or ongoing maintenance may be limited. While LLMs hold promise as tools for disseminating accurate health information, existing general-purpose models often fall short in addressing this specialized domain. They struggle with low accuracy, lack of cultural sensitivity, and the tendency to generate verbose or overly complex responses that fail to resonate with diverse user groups [10]. In recent times, many chatbot-based solutions have emerged such as Flo [11] and SnehAI [12]. These chatbots have advanced menstrual health education by providing personalized cycle tracking, symptom management, and showing efficacy in improving health literacy and user engagement [13]. Despite these advancements, existing chatbots often face limitations. They tend to have shallow conversational interactions, limited flexibility, and difficulty adapting to diverse cultural backgrounds. Additionally, their emphasis largely remains on symptom monitoring and cycle predictions, rather than providing a more comprehensive and empathetic educational experience. These gaps require specialized, culturally adaptive language models capable of delivering thorough menstrual health education interactively.

To tackle these challenges, this study aims to develop and evaluate MenstLLaMA, a specialized LLM tailored for menstrual health education, and to assess its effectiveness across clinical accuracy, cultural sensitivity, and user satisfaction, in comparison to existing general-purpose LLMs. Built upon the LLaMA-3-8B-Instruct model [14] and fine-tuned using our custom MENST dataset comprising 23,820 question-answer pairs, MenstLLaMA bridges critical gaps in existing LLMs. Our comprehensive evaluation demonstrated its superior performance compared to leading models, including GPT-4o [15] and Claude-3 [16], across both automated metrics and clinical expert assessments. To evaluate MenstLLaMA by medical practitioners and to conduct a user study, we further developed ISHA (Intelligent System for Menstrual Health Assistance), an interactive chatbot powered by MenstLLaMA. This study highlights the potential of MenstLLaMA as an empathetic, culturally aware AI companion for advancing menstrual health education in India.

### Related Work

LLMs have shown promise in various healthcare domains; but their application to menstrual health education has been limited. Before initiating this study, we conducted a systematic review of AI applications in menstrual health education, searching PubMed, Google Scholar, and arXiv for articles published between 2020 and 2024 using the keywords “artificial intelligence,” “language models,” “menstrual health,” and “health education.” While general-purpose LLMs like GPT-4 and Claude-3 have proven effective in medical knowledge tasks, no specialized models exist for menstrual health education. However, these general purpose models are prone to hallucinate, i.e., generating fluent yet factually incorrect medical content [17]. This can lead to severe compromise to the safety and trustworthiness in healthcare applications [18]. This risk is critical in sensitive domains like menstrual health, where inaccurate advice can lead to misinformation and potential harm. Previous efforts leveraging AI in menstrual health have primarily focused on period-tracking applications and symptom prediction, with limited emphasis on comprehensive menstrual health education or counseling supported by specialized LLMs. Providing accurate and accessible menstrual health information at scale poses a significant challenge in healthcare systems worldwide. Resource constraints and societal barriers limit traditional approaches relying on healthcare professionals and educational programs. Recent advances in AI and natural language processing offer promising solutions for delivering personalized menstrual health education. Nonetheless, general-purpose LLMs often struggle with cultural insensitivity, providing responses which are misaligned with local norms and socio-cultural taboos on menstruation [19]. Such limitations can hinder effective communication in culturally nuanced contexts, highlighting the need for culturally aware and domain-specific models. However, research on specialized language models for sensitive healthcare domains remains limited, especially in culturally nuanced areas like menstrual health. Most studies on menstrual-related domains are centred around chatbots. For instance, Cunningham et al. [13] used a chatbot, Flo [11], a period and reproductive health tracker providing personalized menstruation/ovulation predictions, symptom forecasts, expert-reviewed content, and an anonymous community platform. They used Flo to measure its efficacy in improving menstrual health literacy and well-being outcomes with and without PMS and PMDD. Kim et al. [20] conducted a randomized controlled trial demonstrating that co-designed mobile educational modules significantly improved sexual knowledge (video) and menstrual knowledge (PDF), with comparable overall effectiveness between formats. Another study reports the development of an AI-based reproductive and menstrual health learning module and found it effective as an educational tool, addressing cognitive accessibility through revised materials and interactive design [21]. The research highlighted the module’s feasibility after validation by students, teachers, and experts. While mobile applications for menstrual tracking have shown promise in facilitating real-time monitoring of menstrual health, recent studies indicate that app users and non-trackers share comparable demographic and menstrual cycle characteristics, suggesting the potential broad applicability of digital interventions in this domain [22]. This is evident with chatbots such as SnehAI, which has effectively addressed sensitive health topics in India by successfully engaging millions of users in discussions about taboo subjects while providing accurate and culturally appropriate information [12]. Apart from the chatbots, study also reveals that web-based menstrual health resources can effectively improve health literacy and encourage medical consultation among young people [23]. However, these platforms typically offer limited interactivity.

Additionally, educational interventions have shown promise in improving menstrual health knowledge. For instance, a study in Iran demonstrated that structured health education programs significantly improved menstrual health knowledge among adolescent girls, highlighting the effectiveness of targeted educational approaches [24]. Sosnowski et al. [25] proposed a hierarchical three-layer architecture that analyzes menstrual cycle features to predict ovulation dates and detect health risks like Premenstrual Syndrome and Luteal Phase Defect. While this approach demonstrated success in cycle prediction and risk assessment, it focused primarily on physiological tracking rather than comprehensive menstrual health education.

## Methods

### Study Design and Setting

This study followed a structured approach, encompassing three main stages: dataset creation, model development, and evaluation. The first stage involved the creation of the MENST dataset, a comprehensive collection of 23,820 question-answer pairs covering various facets of menstrual health. This dataset formed the foundation for developing and evaluating MenstLLaMA, a novel generative AI model built upon the LLaMA-3 architecture [11].

In the model development phase, we fine-tuned the LLaMA-3 model using the MENST dataset to improve its capacity to generate accurate, culturally sensitive, and empathetic responses for menstrual health education. To assess MenstLLaMA’s performance, we conducted a comparative analysis involving other state-of-the-art GenAI models (e.g., GPT-4o and Claude-3) and evaluated its effectiveness against expert clinical assessments.

Model fine-tuning and analysis were performed using Python-based machine learning frameworks, including PyTorch and Hugging Face’s Transformers library [26]. The fine-tuning process leveraged high-performance computing resources to enhance model performance and efficiency. We employed the SciPy [27] and NLTK [28] libraries for comprehensive statistical analysis, calculating various evaluation metrics to ensure reproducibility and robustness in our assessments. In addition to these computational evaluations, human-based assessments were conducted by clinical experts and end-users. These assessments focused on evaluating the model’s relevance, understandability, and cultural sensitivity, particularly in menstrual health education. This dual approach ensured a thorough and well-rounded evaluation process.

### Dataset Creation

#### Dataset Sources

The MENST dataset was compiled from various reputable sources, including health information portals, medical institutions, government websites, global organizations, and educational platforms. Most of the dataset was sourced from official medical documents, where we extracted and curated relevant FAQs. Additionally, we incorporated an existing question-answer dataset on menstrual health, specifically the Menstrual Health Awareness Dataset [29]. This dataset, consisting of 1,986 question-answer pairs, and all the collected QA pairs were manually annotated with metadata (see *Metadata creation* for the detailed metadata description).

To enrich the dataset further, we employed prompting techniques with advanced language models like GPT-4 and Gemini 1.5 Pro, generating additional question-answer pairs from relevant menstrual health documents. This process helped to create a robust and comprehensive dataset tailored to menstrual health education, ensuring accuracy, cultural relevance, and empathy in the content. In the following subsections, we provide a detailed breakdown of the dataset’s structure, metadata creation, and the paraphrasing strategies employed to augment its coverage.

#### Metadata Creation

We created comprehensive metadata for all the documents to facilitate efficient data management and detailed catalogues of menstrual health topics across various demographics and contexts. We tagged each document with a unique document ID starting with ‘D’ or ‘F,’ where unstructured documents or paragraphs were prefixed with ‘D’, which stands for documents. In contrast, ‘F’ or Frequently Asked Questions (FAQs) was prefixed to the already structured or question-answer pair documents. In addition, we included the source and link of each document, along with the topics covered by the document, as part of the metadata.

The dataset must be structured as question-answer pairs to fine-tune an LLM for tasks such as question-answering or conversational interactions. To achieve this, we processed unstructured documents using LLMs to generate corresponding question-answer pairs. The preprocessing methodology for transforming unstructured documents into question-answer format is detailed in the following section. These LLM-generated question-answer pairs were combined with pre-existing pairs to construct the final MENST dataset. Table 1 outlines the metadata elements for the original collected documents, while Table 2 provides the metadata elements for the final processed question-answer dataset.

**Table 1:** Metadata schema for source documents in the MENST dataset.

| Metadata Labels | Description |
| --- | --- |
| Document ID | Each document is assigned a unique ID, starting with ‘D’ or ‘F’. This naming convention helps distinguish documents with only paragraph texts and question answers. |
| Document Name | Name of the document, such as title or main heading. |
| Source | Source of the document containing the name of the website or organisation. |
| Link | URL taking to the document |
| Topic | The topic on which the document is based, for example, is irregular menstruation. |

**Table 2:**
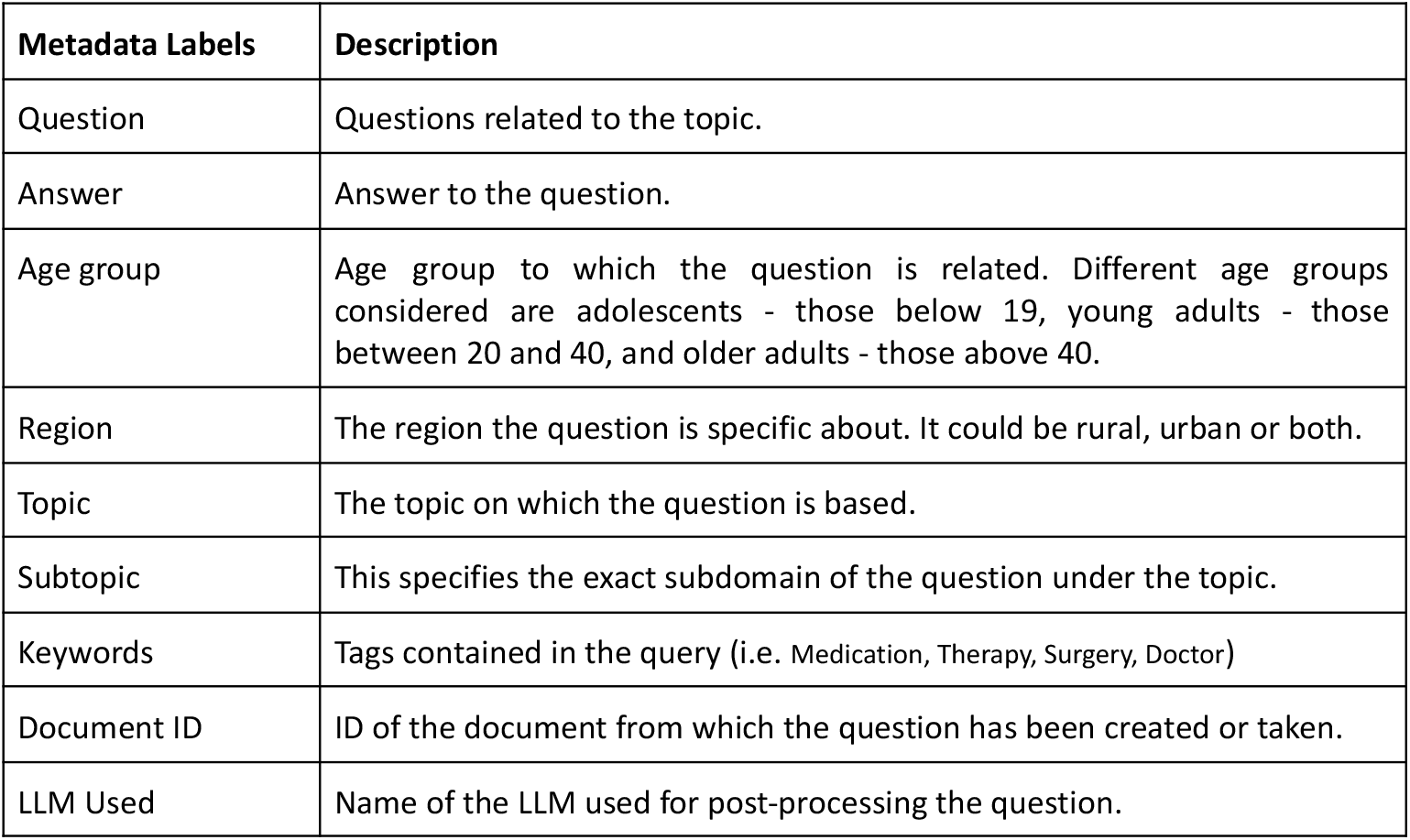
Metadata schema for question-answer pairs in the MENST dataset.

| Metadata Labels | Description |
| --- | --- |
| Question | Questions related to the topic. |
| Answer | Answer to the question. |
| Age group | Age group to which the question is related. Different age groups considered are adolescents - those below 19, young adults - those between 20 and 40, and older adults - those above 40. |
| Region | The region the question is specific about. It could be rural, urban or both. |
| Topic | The topic on which the question is based. |
| Subtopic | This specifies the exact subdomain of the question under the topic. |
| Keywords | Tags contained in the query (i.e. Medication, Therapy, Surgery, Doctor) |
| Document ID | ID of the document from which the question has been created or taken. |
| LLM Used | Name of the LLM used for post-processing the question. |

Collaborating with expert gynaecologists, we created a taxonomy for the topics covered in the menstrual health documents. Primary categories include *Anatomy, Normal Menstruation, Abnormal Menstruation, Pregnancy, Lifestyle, Support*, and *Society*. Each primary category was further subdivided into specific subtopics. For instance, *Normal Menstruation* includes subtopics like *Menarche, Menopause, Normal Flow*, and *Normal Cycle*, while *Abnormal Menstruation* covers issues such as *Abnormal Bleeding, Irregular Periods, Menstrual Pain, PCOS (Polycystic Ovary Syndrome)*, and *PMS (Premenstrual Syndrome)*. A detailed breakdown of the dataset’s taxonomy is provided in Figure 1.

**Figure 1:**
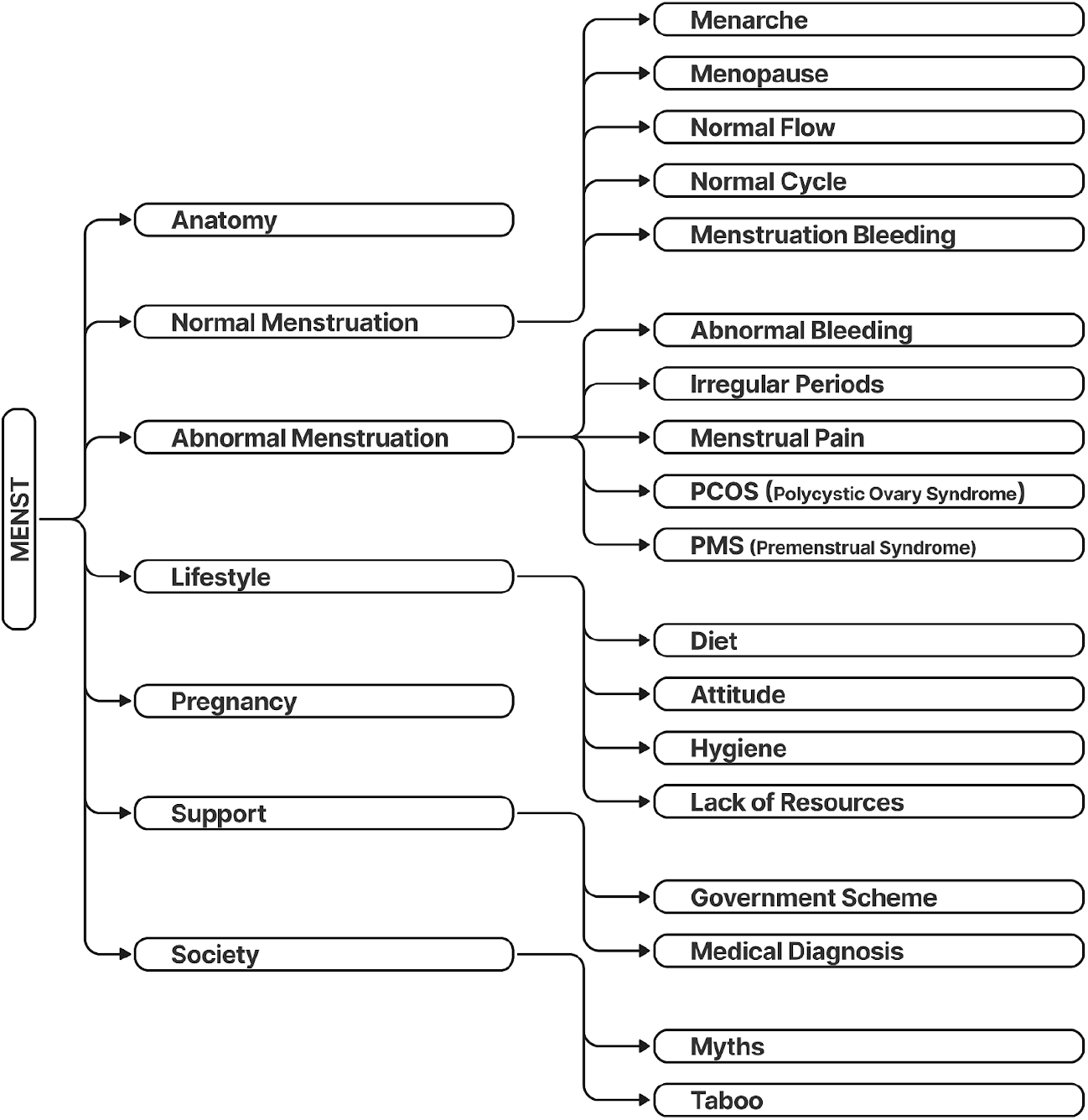
Taxonomy of menstrual health topics in the MENST dataset. The taxonomy consists of seven primary categories (Anatomy, Normal Menstruation, Abnormal Menstruation, Pregnancy, Lifestyle, Support, and Society) with corresponding subtopics as described in the Metadata section.

#### Question-Answer Pair Creation

Our primary dataset included 88 documents, of which 13 were question-answer documents (FAQ). These FAQ documents, sourced from official medical portals, were used as a gold test set (Set-1). For the remaining 75 unstructured documents, we used GPT-4 and Gemini 1.5 Pro to generate question-answer (QA) pairs. These generated pairs were then validated by domain experts.

We employed a standardized prompt template to ensure the consistency of the QA generation process. The prompt template used three random sample QA pairs from Set-1 to provide context for the language models, as shown in Figure 2.

**Figure 2:**
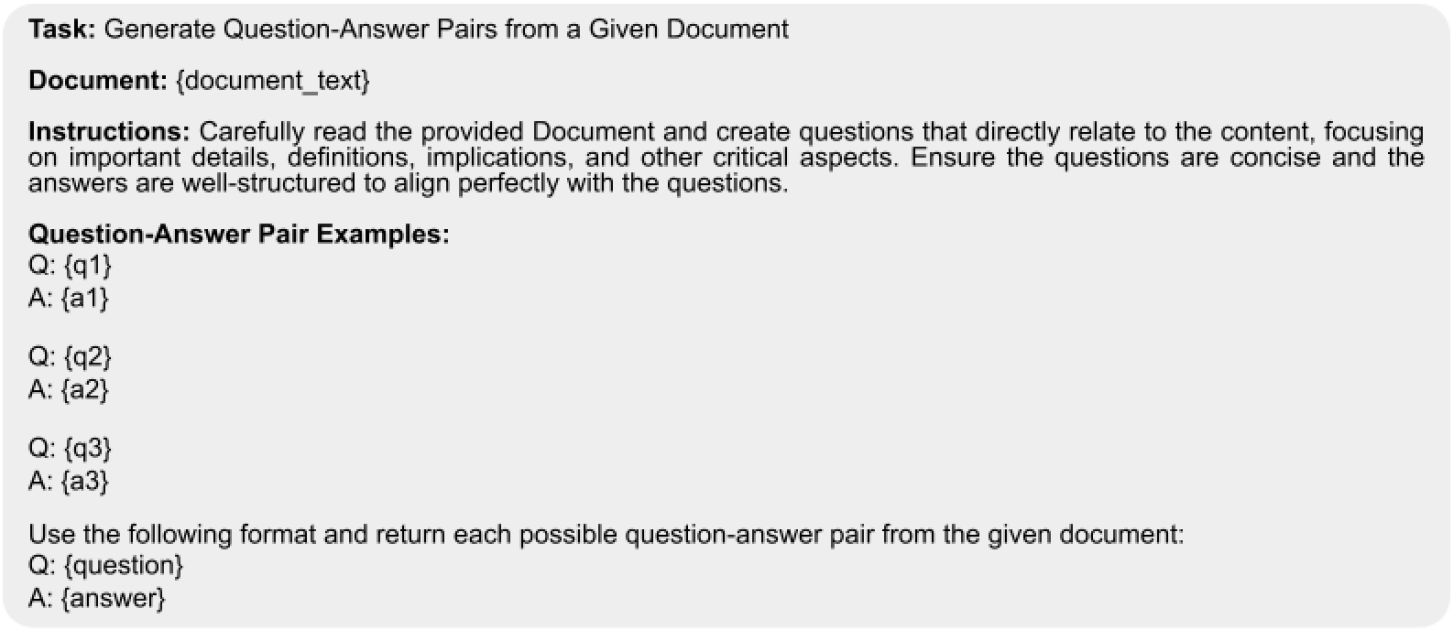
Prompt template structure for curating question-answer pairs from unstructured medical documents. The template includes four components – (1) a task description specifying the generation of relevant questions from document content, (2) the unstructured document from which the QA pair is to be generated, (3) instructions for creating concise, well-structured questions focusing on critical aspects, and (4) example format showing three QA pairs from gold standard data to guide the generation process.

#### Paraphrasing for Dataset Augmentation

To enhance the diversity of the dataset, we employed paraphrasing techniques. This approach improves the model’s handling of varied user expressions and contexts. By prompting GPT-3.5 with instructions shown in Figure 3, we implemented four distinct paraphrasing strategies:

1. **Scenario-based paraphrasing** helps questions include real-life situations, making the model’s learning data more responsive to human input. For example, “Can I cook while I am on my period?” could be paraphrased as “Today while I was in the kitchen, my mother scolded me as I entered the kitchen on my period. Is it a sin to cook during this phase?” This helps the model better understand the queried question and respond in ways more aligned with human conversational patterns.
2. Rephrasing from a **male point of view** gives a male’s perspective on the question, often lacking direct experience with menstruation. This perspective is essential as it expands the model’s understanding and ability to engage with questions from individuals indirectly affected by the issue. For example, “I am experiencing painful periods. Is this normal?” becomes “My wife is experiencing painful periods. Is this normal?”. This ensures that the model can adequately address queries from a broader audience.
3. **Changing the sentence structure** introduces linguistic diversity in the question, which is crucial for developing syntactic flexibility in a model. For example, “Is it normal to start menstruating at the age of 12?” can be rephrased as “I am 12 years old and just got my first period. Is this okay?”. This diversity in data helps the model learn the flexibility needed to understand personalized questions.
4. Paraphrasing from the **rural women’s perspective** captures the language and concerns of women in rural settings, who may have different cultural and educational backgrounds. For example, “I don’t have access to sanitary pads. What can I use instead?” is paraphrased as “In my village, there are no sanitary pads. What can I use instead?” This helps the model respond appropriately to a diverse demographic, especially village students.

**Figure 3:**
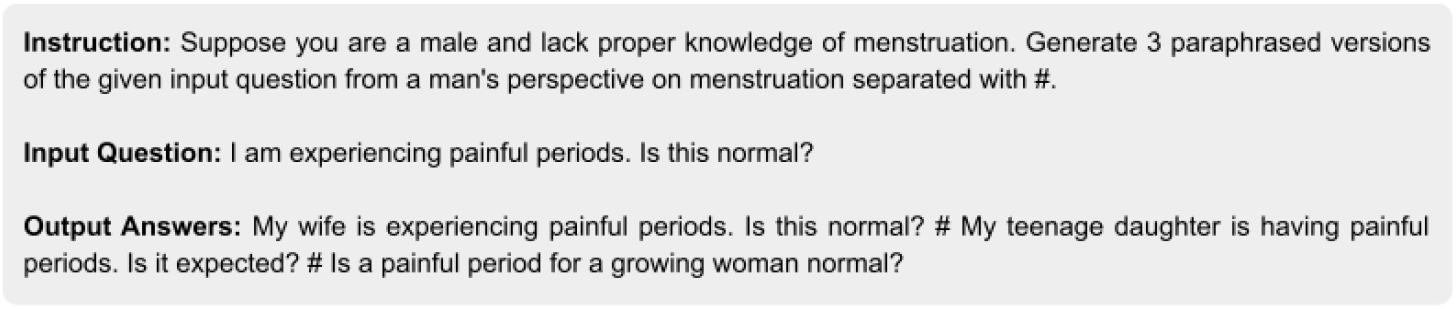
Example of the prompt template used for dataset augmentation using paraphrasing the questions. The template demonstrates the male perspective paraphrasing strategy, showing (1) the input instruction that specifies the paraphrasing task from a male’s perspective, (2) a sample input question about menstrual pain, and (3) three generated paraphrased versions that maintain clinical accuracy while incorporating different male familial relationships (wife, teenage daughter, growing woman). This template generated diverse yet contextually relevant question variations for the MENST dataset, enhancing the MenstLLaMA’s ability to handle queries from different perspectives and social contexts.

Each question from the raw dataset was paraphrased across four distinct scenarios, resulting in 23,820 question-answer pairs. A breakdown of the dataset’s structure is provided in Table 3.

**Table 3:**
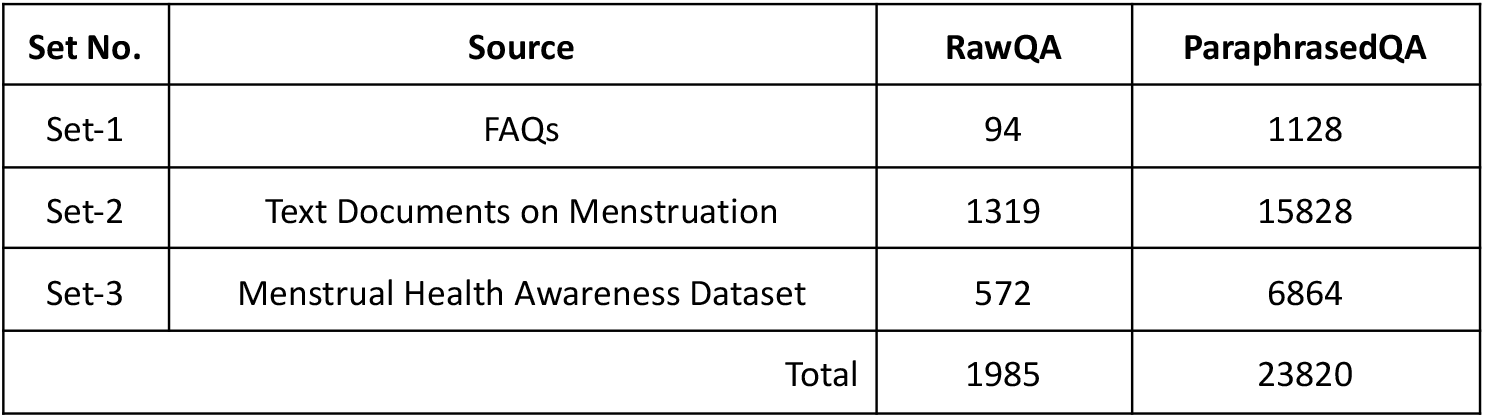
Distribution of QA pairs in the MENST dataset across different data sources before and after paraphrasing augmentation.

| Set No. | Source | RawQA | ParaphrasedQA |
| --- | --- | --- | --- |
| Set-1 | FAQs | 94 | 1128 |
| Set-2 | Text Documents on Menstruation | 1319 | 15828 |
| Set-3 | Menstrual Health Awareness Dataset | 572 | 6864 |
| Total |  | 1985 | 23820 |

#### Expert-Compiled Test Dataset

Collaborating with medical professionals, we developed an expert-curated test dataset to evaluate the model. We asked experts to compile questions they commonly encounter in clinical practice and provide appropriate answers based solely on their clinical expertise, without reference to external materials. Using our metadata framework (7 topics, 19 subtopics), we obtained 200 QA pairs reflecting real-life clinical scenarios. This expert-curated set was used as a gold test set for evaluating the model’s performance.

### Model Development

We developed MenstLLaMA, a specialized language model for menstrual health, by fine-tuning the Meta-LLaMA-3-8B-Instruct model [14] on our curated MENST dataset, which comprises 23,820 question-answer pairs. Fine-tuning was conducted using a parameter-efficient fine-tuning (PEFT) strategy [30], with a specific focus on Low-Rank Adaptation (LoRA) [31] to align the model with the MENST dataset. PEFT is an adapter-based fine-tuning approach that optimizes training by adjusting only a small subset of parameters while keeping the majority of the base model frozen. LoRA, in particular, introduces a low-rank matrix into the attention mechanism, enabling the model to effectively learn task-specific patterns with reduced computational overhead. Alternative strategies such as full fine-tuning, other adapter-based methods (e.g., prefix-tuning, prompt-tuning), and retrieval-augmented generation (RAG) were also considered. Full fine-tuning is resource-intensive and less practical for iterative experimentation. RAG introduces a dependency on external retrieval systems, which may not always yield culturally sensitive or domain-relevant content. While other adapter-based methods are lightweight, LoRA offers a favorable trade-off between computational efficiency, performance, and ease of integration into transformer architectures. Thus, LoRA was selected as the most suitable approach for our domain-adaptive fine-tuning of MenstLLaMA. This approach allows for efficient adaptation across diverse conversational contexts while maintaining performance comparable to full fine-tuning.

Instruction fine-tuning was performed on the LLaMA-3-8B-Instruct model using a structured prompt format to distinguish between instructions and responses. Each QA pair was reformatted, as shown in Figure 4. This format ensured that the model could clearly distinguish between instructions and responses. This improved its ability to handle complex context-sensitive conversations by enabling the model to effectively capture the nuances of dialogue.

**Figure 4:**
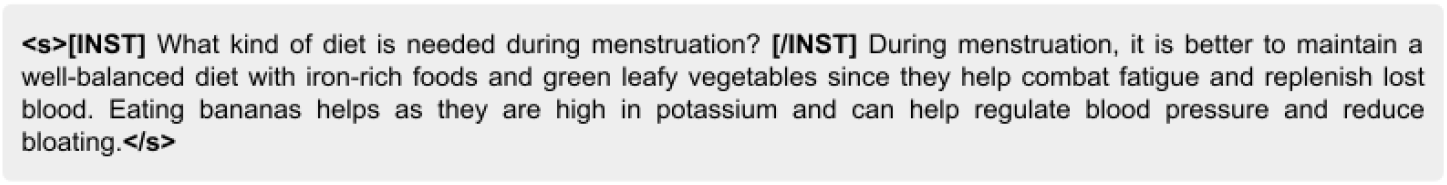
Instruction fine-tuning format for MenstLLaMA, illustrating the standardized structure for QA pairs. The format demonstrates the conversion of the QA pairs into LLaMA instruction syntax (**<s>[INST]** *question* **[/INST]** *answer* **<s>**). This example illustrates a dietary question during menstruation, with the response structured to provide clear, scientific information about nutritional needs. The format enables the model to distinguish between the question and their corresponding responses.

We used Meta-Llama-3-8B-instruct [32], an 8 Billion parameter model, as the base model for our model configuration. We finetuned the model with a maximum sequence length of 2048 tokens and leveraged LoRA for efficient training. We applied 4-bit quantization using the NF4 type to optimise memory usage. We used Paged AdamW [33] as the optimizer with 32-bit precision for efficient gradient updates. The training hyperparameters were fine-tuned to balance learning rate, batch size, and gradient accumulation steps, allowing us to achieve optimal performance with minimal computational overhead. The training process involved five epochs with a learning rate of 2e-4, a warmup ratio of 0.03 and a maximum gradient norm of 0.3. We utilised the NVIDIA A100 GPU with 80GB of memory for the finetuning process. Notably, we leveraged free-tier API credits offered by OpenAI and Google as part of their academic access programs, ensuring zero financial cost for this phase. The total time taken for dataset curation, prompt engineering, and API-based generation was approximately 10 weeks. Since the finetuning (model training) and model deployment at the time of user evaluation was done on local hardware (NVIDIA A100 GPU), there were no associated cloud infrastructure or compute rental costs. As such, the overall process incurred no additional financial expenditure.

#### Development of an Interactive Chatbot

To enable qualitative evaluations by medical practitioners and facilitate case studies, we developed an interactive chatbot powered by the fine-tuned MenstLLaMA model, ISHA (**I**ntelligent **S**ystem for Menstrual **H**ealth **A**ssistance). ISHA was designed to ensure a seamless conversational flow by leveraging contextual information from previous utterances within the same session. During the fine-tuning and inference stages, we incorporated tailored instructions into the model’s prompts to enhance the chatbot’s empathy and emotional awareness. This approach aimed to create a contextually accurate chatbot that is emotionally attuned to the nuances of the menstrual health domain. All interactions with ISHA were conducted in a controlled research environment without logging personal identifiers, and strict data handling protocols were followed to ensure user privacy and security. Although the ISHA interface is not currently hosted for public access due to infrastructure and maintenance costs, the underlying MenstLLaMA model and supporting codebase have been made publicly available. Figure 5 presents the ISHA interface.

**Figure 5:**
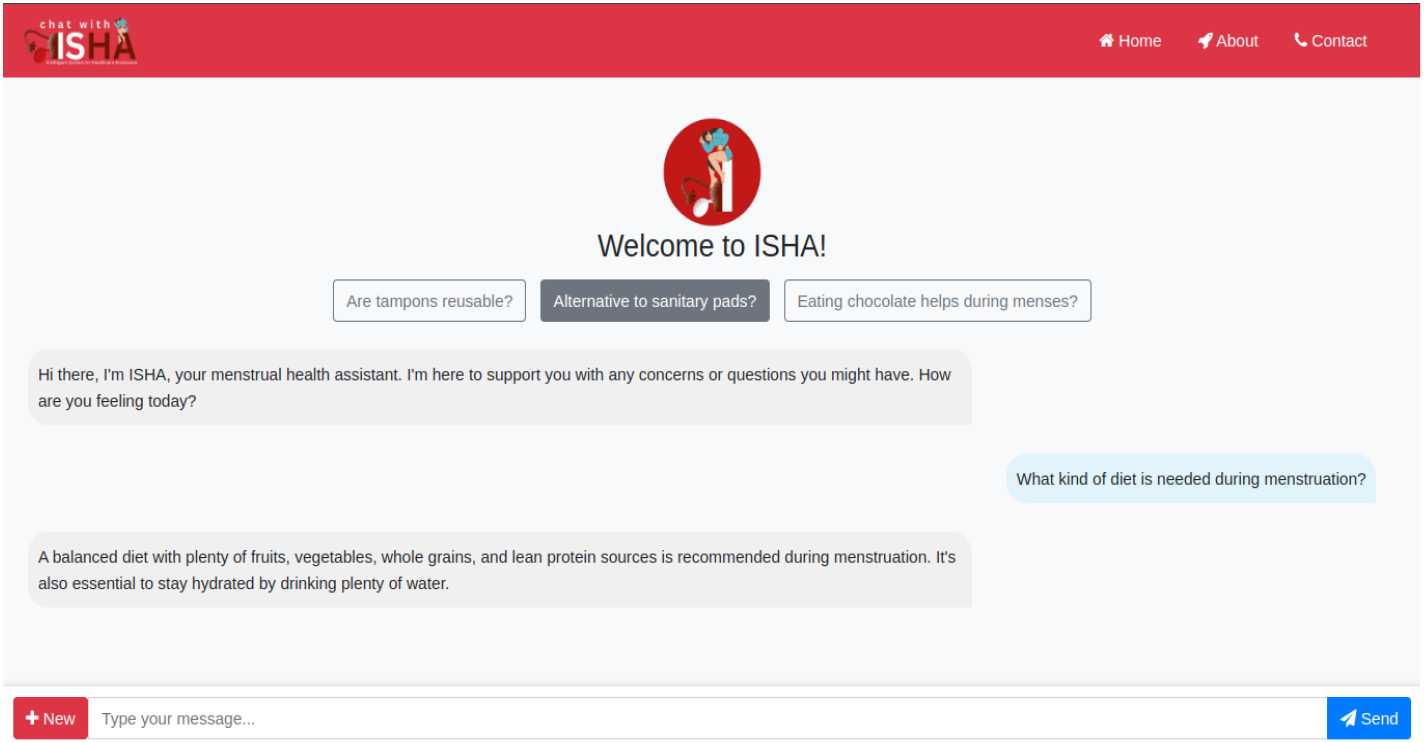
Interface of the ISHA chatbot powered by MenstLLaMA.

### Baseline Models

We compared MenstLLaMA against nine state-of-the-art LLMs, including three closed-source models (GPT-4 [12], Gemini 1.5 Pro [34], Claude-3 [13]) and six open-source models (Mistral [35], LLaMA-3 [11], GPT-2 [36], Orca2 [37], Falcon [38], and Phi3 [39]).

#### Evaluation

The evaluation of MenstLLaMA and other state-of-the-art closed and open-sourced models followed a comprehensive four-step approach to assess the model’s performance across technical accuracy, clinical relevance, and user satisfaction. These steps included **automatic evaluation** using standard NLP metrics, **clinical expert evaluation, medical practitioner feedback** in a simulated setting, and a **user-centred evaluation**. This multi-faceted approach provided a balanced understanding of the model’s strengths and limitations, particularly in the sensitive domain of menstrual healthcare.

#### Automatic Evaluation

MenstLLaMA was benchmarked against leading conversational models, including proprietary systems (GPT-4o, Gemini 1.5 Pro, Claude-3) and open-source models (Mistral, LLaMA-3, GPT-2, Orca2, Falcon, Phi3). Given the generative nature of the task, we employed standard Natural Language Processing (NLP) evaluation metrics such as BLEU [40], METEOR [41], ROUGE-L [42], and BERTScore [43] to assess the models against the expert-compiled gold test set, which together evaluated lexical overlap, fluency, semantic similarity, and factual consistency. While BLEU has known limitations in capturing factual alignment and dialogue coherence, we included it to ensure comparability with prior work. We complemented it with newer metrics like BERTScore that better account for semantic nuance and information preservation While BLEU, METEOR, and ROUGE-L provide insight into word and sequence overlap, they are more appropriate for rigid, template-based responses. In contrast, healthcare dialogue often includes semantically valid variations that may not align lexically. In this context, BERTScore offers a more robust evaluation by capturing the semantic similarity of model outputs to expert references.

Therefore, BERTScore serves as a particularly suitable metric in our domain, and we give it greater interpretive weight when comparing model performance. These metrics assessed reference responses’ fluency, relevance, overlap, and semantic similarity. More details on the metrics are presented in Table 4. Following this evaluation, the top four models were selected for further analysis. Detailed results from this comparative analysis are presented in the Results section.

**Table 4:**
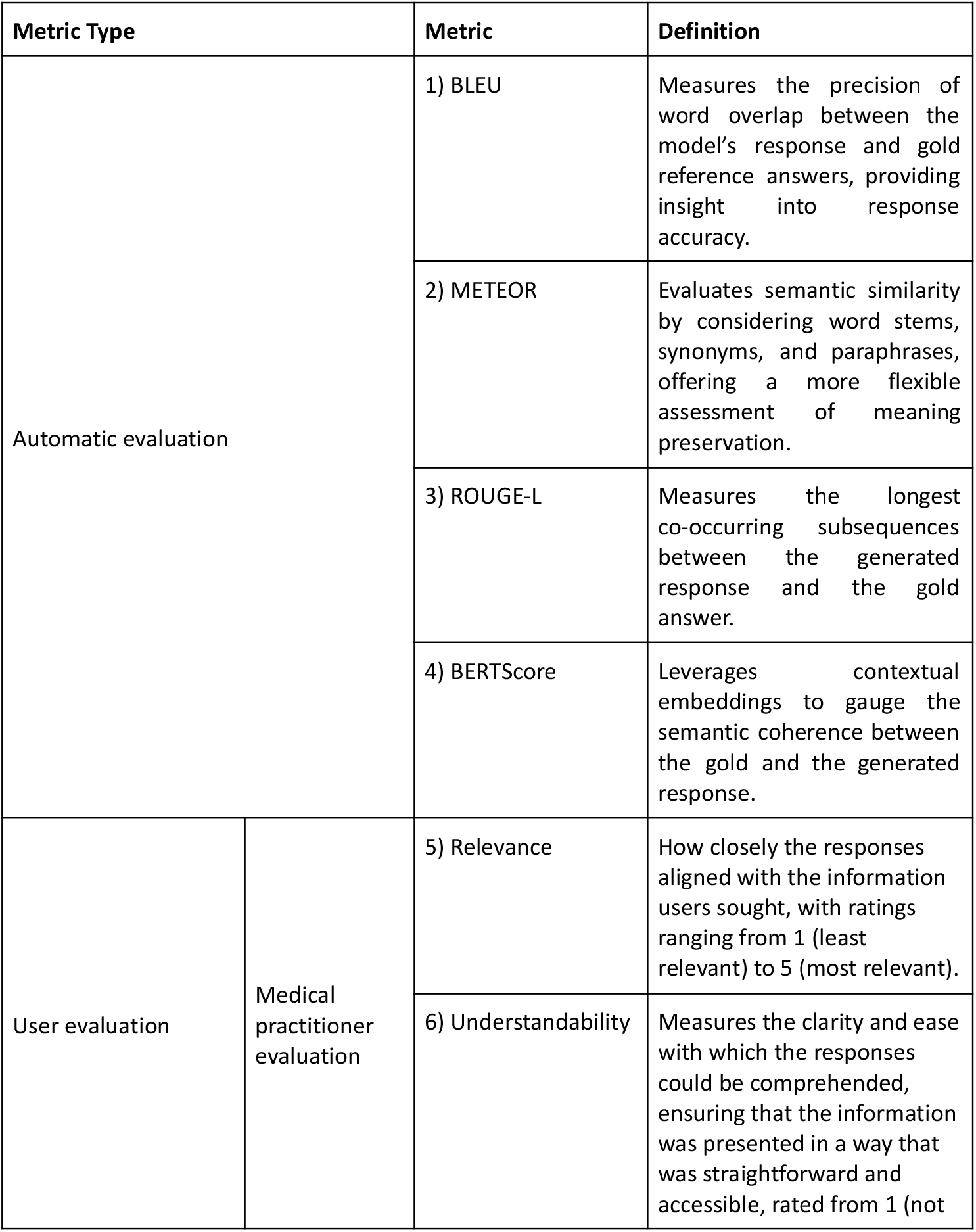
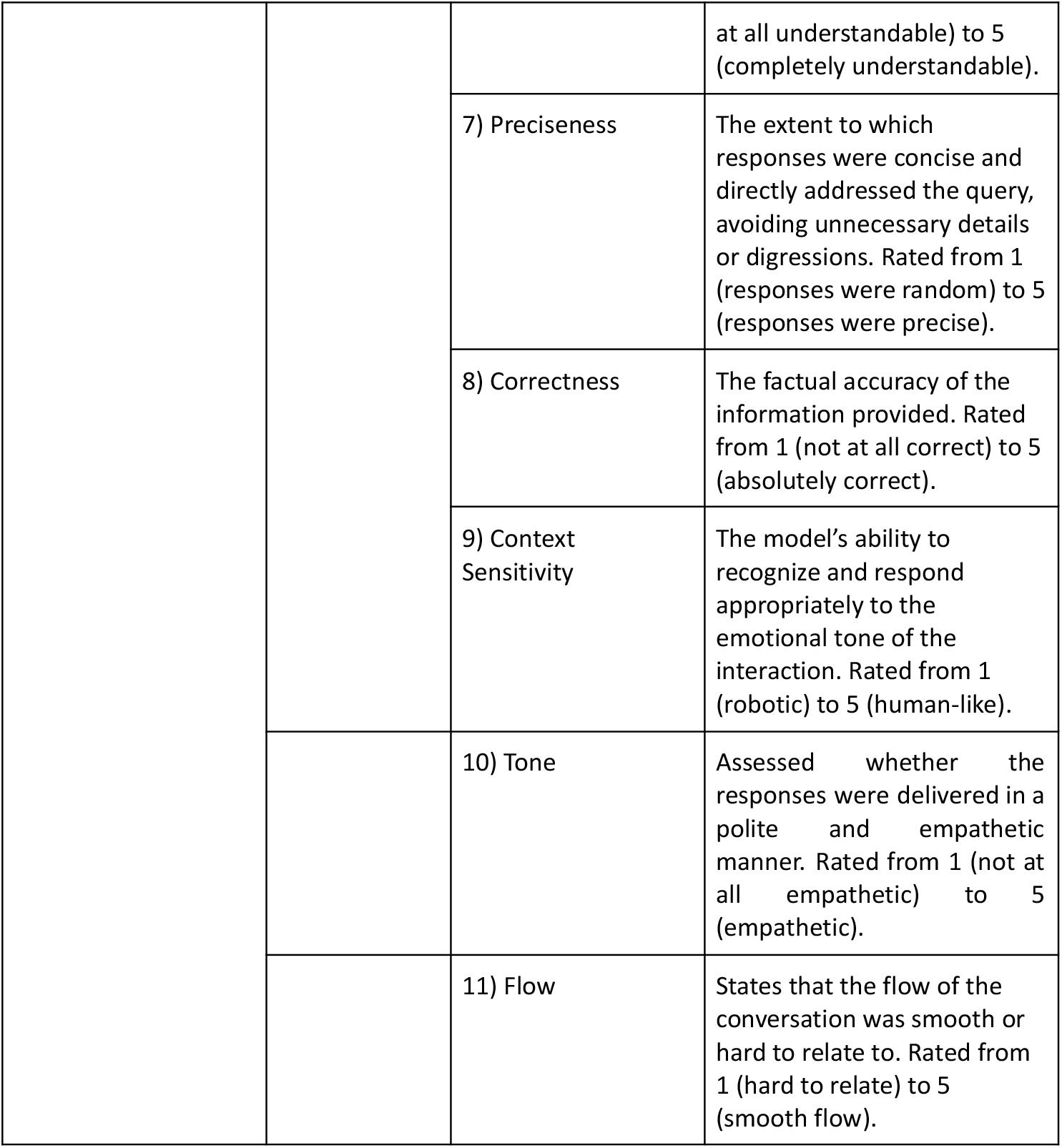
Evaluation metrics, definition and scoring criteria for MenstLLaMA performance. Notes: automatic evaluation metrics (1-4) where all the metrics’ scores range from 0 to 1, with 0 being the least and 1 being the maximum, medical practitioners’ assessment criteria (5-9), and user evaluation criteria (5-11).

#### Evaluation by Clinical Experts

To assess the clinical relevance of our model against the top-performing baseline models based on the automatic evaluation, we engaged six clinical professionals with direct experience in gynecological and sexual health consultations. The panel included three Bachelor of Medicine final-year trainees (n=3) with ongoing clinical rotations in obstetrics and gynecology, two certified physician assistants (n=2) with at least three years of reproductive health experience, and one licensed sexual health therapist (n=1) specializing in adolescent and women’s health education‥ The panel comprised three Bachelor of Medicine trainees (n=3), two physician assistants (n=2), and one sexual health therapist (n=1). These evaluators were recruited via professional networks and academic affiliations and volunteered their time for the study without monetary compensation. These professionals were presented with the 200 questions from the expert-compiled test set, each accompanied by five unlabeled responses: the top four models and one gold-standard response curated by domain experts. The experts selected the most appropriate responses from a set of five, where four responses were generated by the top four models, and the remaining response was sourced from the expert-compiled test set. To ensure objectivity, the sources of the responses were not disclosed to the evaluators, and selections were made solely based on perceived clinical appropriateness and clarity.

#### Evaluation by Medical Practitioners

Following the model comparisons, we further extended the evaluation by asking practicing gynecological professionals to assess MenstLLaMA’s responses in a simulated clinical setting. Using the ISHA chatbot interface, each evaluator was asked to simulate patient interactions and then rate MenstLLaMA’s responses using a set of qualitative metrics. Twelve medical practitioners were recruited – five experienced doctors (n = 5) and seven postgraduate trainees in medicine (n = 7), all regularly interacting with patients in their clinical routines. Recruitment for the study took place through informal, word-of-mouth referrals within healthcare networks. We relied on personal and professional connections to share information about the study, which helped us reach potential participants in a more organic and trusted way. As with the expert reviewers, these practitioners participated voluntarily without financial incentives. We compiled these metrics to assess various aspects of the model’s performance in a clinical context. Each metric was rated on a five-point Likert scale, where 1 signifies the lowest score and 5 the highest. More details on these metrics are reported in Table 4. In this way, we evaluated MenstLLaMA with the experts’ involvement in a simulated clinical setting, providing insights beyond quantitative measures. The results of this evaluation are discussed in the Results section.

#### User Evaluation

For a more comprehensive evaluation of the MenstLLaMA, a user study was conducted with 200 participants, comprising 181 females (n=181) and 19 males (n=19), representing a diverse range of socio-demographic and professional backgrounds. The participants ranged in age from 18 to 52 years (M = 28.4, SD = 7.3) – approximately 72% identified as urban residents, 23% semi-urban, and 5% rural. Educational backgrounds included high-school graduates (7%), undergraduate (49%), postgraduate (31%), and doctoral or equivalent professional qualifications (13%). The cohort included engineering and medical students, researchers, practising doctors, allied health professionals, and industry professionals from the technology, finance, and consulting sectors. The study was conducted primarily across India, Bangladesh, Nepal and Bhutan, thereby integrating regional and cultural diversity into the evaluation process.

The participants were recruited through open calls circulated on university bulletin boards, professional forums, and social media, intending to include a broad spectrum of users. All participants volunteered for the study and were not compensated, which we acknowledge may have introduced interest-based self-selection bias. Prior to participation, informed consent was obtained via a consent modal displayed through the ISHA web interface, where users were briefed about data usage for research purposes. No personal or identifiable information was collected. The participants were instructed to interact with MenstLLaMA through the ISHA chatbot interface and provide user feedback. The participants were asked to evaluate the model’s response based on the same metrics (see Table 4 for the full set of metrics) as those used in the above evaluation by medical practitioners in simulated evaluation, with two additional metrics: *Tone* and *Flow*. These were especially significant in menstrual and gynecological health, where sensitivity and compassionate communication are critical. This pilot study was intended to provide initial feedback on the system’s relevance and accessibility, rather than to reflect broader population-level adoption. Insights from this study will inform future iterations and deployment planning, including projections for user reach and usage scale. These additional metrics were particularly significant in menstrual and gynaecological health, where sensitivity and compassion are paramount. The results of this user study are presented in the Results section.

## Results

We evaluated the MenstLLaMA against the baseline LLMs using both automatic and human-based approaches.

Table 5 reports the automatic scores for the models computed using BLEU, METEOR, ROUGE-L, and BERTScore. In our experimentation, we conducted various prompting strategies for the baseline LLMs viz, zero-shot (no example given in the prompt), one-shot (single example) and two-shot (two examples). MenstLLaMA achieved the highest scores in the BLEU (0.059) and BERTScore (0.911) metrics. While Claude-3 and GPT-4o achieved the highest for the METEOR (0.321) and ROUGE-L (0.253) metrics. Interestingly, Gemini1.5-Pro and LLaMA 3 demonstrate reliable but slightly lower scores in comparison, reflecting their capabilities in specific tasks. The newer models like Mistral and Orca 2 provide strong BLEU and METEOR scores but occasionally underperform in ROUGE-L compared to others. Falcon, while showing potential, falls behind across most metrics. In the zero-shot scenario, the closed-source models (GPT-4o, Claude-3 and Gemini1.5) outperformed all the open-source models. Introducing a single example in the one-shot scenario improved most models. Notably, Mistral showed a significant jump in performance, achieving the highest BERTScore (0.905), surpassing even the closed-source models in this one-shot setting. A similar trend is observed in the two-shot setting, where the Mistral showed a gain over the zero-shot and two-shot settings and even outperformed the closed-source models in the two-shot scenario. Overall, the closed-source models consistently performed well across all scenarios. They showed strong performance even in zero-shot settings, indicating robust generalization capabilities. However, with a few shot prompting, some open-source models, such as Mistral and Phi 3, showed remarkable improvements. Mistral, in particular, achieved competitive performance with closed-source models in one-shot and two-shot settings. While most models improved with additional examples, the degree of improvement was not uniform. Open-source models generally gained more from few-shot examples compared to closed-source models. Notably, our proposed model, MenstLLaMA, exhibited strong performance across all metrics without relying on few-shot examples. This indicates that fine-tuning with the MENST dataset has effectively adapted the model resulting in a model for the menstrual health domain.

**Table 5:**
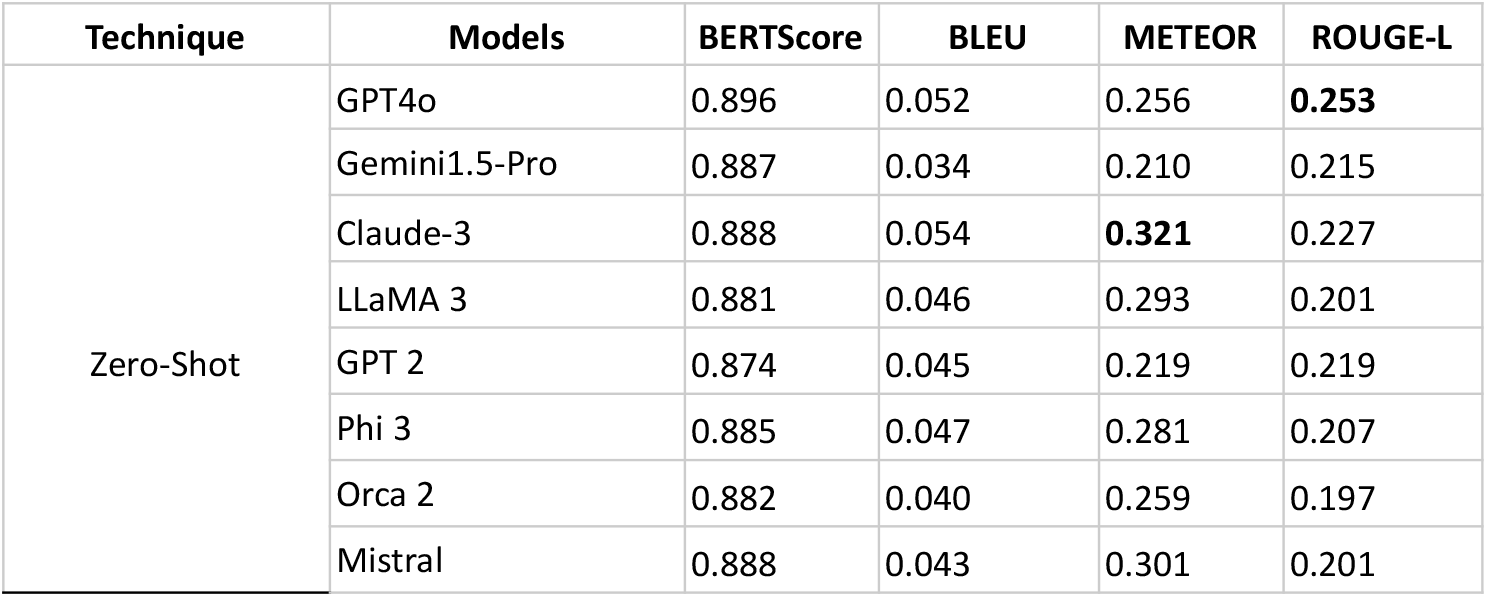
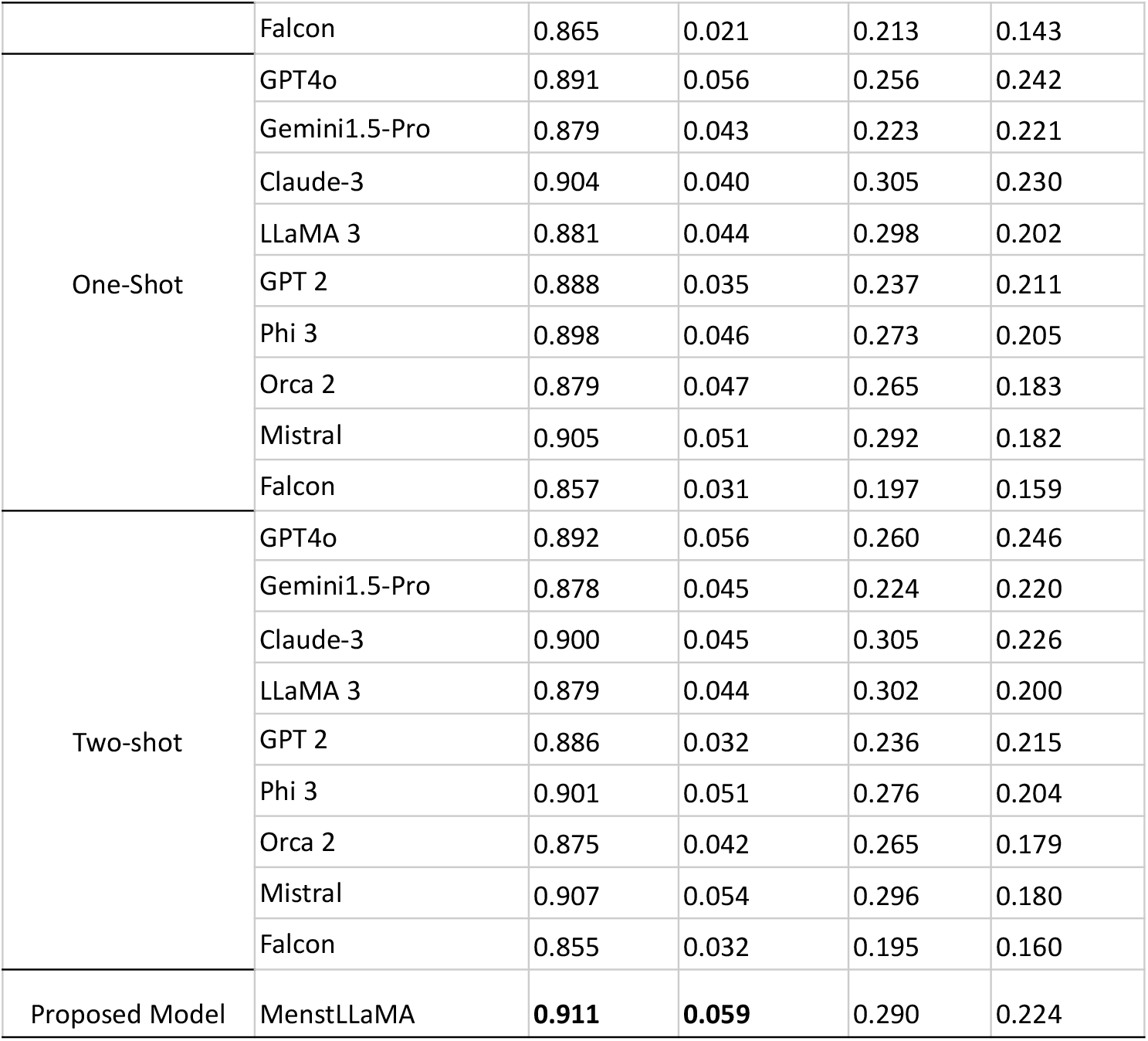
Automatic scores of the models using the metrics: BLEU, METEOR, ROUGE-L, and BERTScore. We employed zero-shot and few-shot (one and two shots) prompting techniques for the baseline models. The scores are computed with respect to the expert-compiled test set.

We selected the top four performing models for further assessment based on these automatic scores: MenstLLaMA, GPT-4o, Claude-3-opus, and Mistral. These models underwent another round of evaluation by the clinical experts on the expert-compiled gold test. To ensure an unbiased assessment, we engaged experts who were not involved in curating the gold test set. MenstLLaMA consistently outperformed other models, including the gold-standard responses, achieving mean expert ratings of 3.97 of *Relevance*, 4.48 of *Understandability*, 3.90 of *Preciseness*, 4.00 of *Correctness* and 3.41 of *Contextual Sensitivity* (all out of 5), with substantial inter-rater agreement (Fleiss’ ϰ ≈ 0.68). This indicates that our model can generate contextually appropriate and medically reliable responses. The fact that MenstLLaMA’s outputs were often preferred over the gold-standard responses shows the model’s effectiveness in interpreting and applying domain-specific knowledge.

Subsequently, we also carried out medical practitioners’ evaluations in a simulated clinical setting using ISHA, MenstLLaMA’s chatbot interface. In this assessment, MenstLLaMA scored 3.5 in *Relevance*, indicating that while the responses were generally pertinent, there were gaps in addressing specific queries fully. For instance, when asked about the safety of using menstrual cups during infections, the model gave a general hygiene tip but failed to mention medical contraindications. *Understandability* was rated at 3.6, reflecting that most responses were clear and easy to comprehend, though explanations involving hormonal regulation sometimes used overly technical language. *Preciseness* scored 3.1, suggesting that although the chatbot provides relevant information, there is some inconsistency in maintaining concise and directly relevant answers. *Correctness* was rated at 3.5, underscoring that while the chatbot delivered accurate responses for common issues, additional updates are needed, particularly regarding recent guidelines (i.e., the model incorrectly cited outdated WHO iron supplementation guidelines in one instance) and less frequently addressed topics. Notably, the model excelled in *Context Sensitivity* with a score of 4, demonstrating a strong ability to respond empathetically and understand the emotional nuances in interactions.

Finally, in the user case study, the users rated the model across seven key metrics on a 5-point scale. The users rated relatively high in *Relevance* (4.3), indicating that users generally found the responses pertinent to their queries. *Understandability* received the highest score (4.7), reflecting the clarity and ease with which users could comprehend the information provided. *Preciseness* was rated 4.28, showing that while responses were generally concise and to the point, there remains a need to fine-tune the output to avoid verbosity and redundancy. *Correctness* was rated 4.1, affirming that users found the chatbot’s information mostly accurate, though some responses could benefit from more specific and detailed updates. The evaluation also highlighted positive scores in *Flow* (4.2) and *Tone* (4.6), with users appreciating the polite and empathetic nature of the interaction. However, *Context Sensitivity* scored lower at 3.9, suggesting room for improvement in how well ISHA adapts to and understands subtle emotional cues in conversation.

## Discussion

### Principal Findings

In this study, we introduced the MENST dataset and MenstLLaMA. The MENST dataset was used to train the MenstLLaMA, a novel open-source large language model designed explicitly for the menstrual health domain. Our comprehensive evaluation highlighted MenstLLaMA’s strong performance across both automated and expert evaluations. It outperformed several state-of-the-art general purpose LLMs (both open and closed source) in automatic and human evaluation metrics, with users and clinicians rating its responses as effectivene in delivering accurate, accessible, and culturally sensitive menstrual health information.

Our domain-specific MenstLLaMA demonstrated competitive performance compared to state-of-the-art general-purpose LLMs, including open-source and closed-source LLMs. Notably, it achieved superior results in automatic evaluation metrics, such as BLEU (0.059) and BERTScore (0.911), without requiring few-shot examples. While closed-source models like GPT-4o and Claude-3 showed robust performance in zero-shot settings, MenstLLaMA’s consistent performance indicates successful adaptation to the menstrual health domain. This underscores the potential of domain-specific fine-tuning to generate fluent, adequate and contextually relevant responses. As a domain-specific model fine-tuned for menstrual health, MenstLLaMA shows consistent quantitative improvements, which is further backed by the improvements in our intensive qualitative evaluation pipeline, thus justifying its development. These qualitative improvements which are critical in sensitive health domains, advocate the need for a specialized model like MenstLLaMA.

Clinical expert evaluations showed encouraging results, with MenstLLaMA’s responses often preferred over gold-standard answers. This preference suggests that the model captures domain knowledge effectively and aligns with clinical expertise. The model’s ability to generate responses that the experts rated more appropriate than the curated answers highlights its potential as a reliable source of menstrual health information. The simulated clinical setting evaluation using the ISHA chatbot revealed both strengths and areas for improvement. The medical practitioners interacted with ISHA, acted like patients, and evaluated the model’s response based on qualitative metrics. While the model demonstrated strong empathetic capabilities (*Context Sensitivity*: 4.0), its performance in *Preciseness* (3.1) suggests room for improvement in generating concise, focused responses. The modest improvement in *Preciseness* reflects MenstLLaMA’s intended role as a supportive, culturally sensitive informational tool rather than a clinical decision making system. Given the sensitivity and taboo surrounding menstrual health in India, the model prioritizes empathetic, elaborative responses to create a safe, private environment for discussing sensitive topics while minimizing hallucinations. This approach naturally affects response length. The findings indicate that while domain-specific training successfully improved the model’s emotional intelligence, further work may be needed to enhance its ability to provide more precise information without compromising empathy or cultural appropriateness.

These findings position MenstLLaMA as a candidate for pilot implementation within digital support services, particularly in regional health initiatives focused on women’s health. A potential next step involves trialing an enhanced version of MenstLLaMA in collaboration with community health workers or digital health platforms in South Asia, where menstrual health education is both urgent and underserved.

User evaluation results were particularly promising for real-world applications. High scores in understandability (4.7) and relevance (4.3) suggest that MenstLLaMA effectively bridges the gap between clinical accuracy and user comprehension. The strong performance in tone (4.6) and flow (4.2) demonstrates the model’s ability to maintain engaging, empathetic conversations while generating accurate and relevant health information.

To this end, the strength of MenstLLaMA lies not only in its technical performance but in its capacity to deliver context-sensitive health education at scale. Its ability to offer culturally tailored, empathetic responses opens new possibilities for reaching populations who may not engage with traditional menstrual health education due to stigma or lack of access. These findings suggest that specialized LLMs like MenstLLaMA can act as an accessible bridge between formal healthcare systems and community needs, particularly in low-resource settings. Moreover, the model’s preference over gold-standard answers in expert evaluation highlights the potential of data-driven approaches to complement and support clinician-driven education strategies.

## Limitations

Our study has several limitations. While the MENST dataset covers a broad range of topics, it does not fully represent all cultural perspectives or demographics, and although diverse user groups were included in our evaluation, longitudinal studies are needed to assess sustained impact. The model’s current text-based interface may limit accessibility for users with varied communication needs or digital literacy. As menstrual health norms evolve, periodic updates are necessary to maintain relevance. Like all LLMs, MenstLLaMA is prone to hallucinations and plausible but incorrect outputs, which can pose risks in health contexts, despite safeguards such as prompt design and fine-tuning. There is also potential for ethical misuse if deployed without oversight, emphasizing the need for human-in-the-loop supervision and responsible implementation. Additionally, there may be inadvertent biases in the model outputs, stemming from the training data and evaluator profiles. Since participation was voluntary and likely attracted individuals already interested in technology or menstrual health, selection bias may also have influenced user evaluation outcomes. While our model has shown promise in controlled evaluations, scaling it for real-world use would involve challenges such as expand demographic representation, improve context sensitivity, integrate the model with broader healthcare interventions, and address real-world deployment challenges such as system integration, digital literacy gaps, trust, and data privacy across diverse user bases.

## Conclusions

MenstLLaMA represents a significant advancement in applying artificial intelligence to menstrual health education. Its performance across multiple evaluation metrics demonstrates that domain-specific language models can effectively bridge the gap between clinical accuracy and accessible health communication. The model’s strong performance in technical metrics and user evaluations suggests its potential as a valuable tool for addressing the critical need for accurate, culturally sensitive menstrual health information. While there are areas for improvement, particularly in precision and context sensitivity, the overall results indicate that MenstLLaMA successfully achieves its primary goal of making menstrual health education more accessible and understandable. As we continue to refine and expand this approach, such specialized language models could play an increasingly important role in improving access to quality menstrual health information and reducing associated stigma globally. Future work will explore responsible deployment pathways, including partnerships with healthcare organizations to pilot MenstLLaMA in targeted regional settings.

## Data Availability

The data supporting the findings of this study are publicly available at Hugging Face. The dataset can be accessed at https://huggingface.co/datasets/proadhikary/MENST.

## Contributors

The contributions of individual authors are as follows: Conceptualisation: TC; Data curation: IM, PKA, MJ, KP, GO; Formal analysis: PKA, IM, SMS; Funding acquisition: TC; Investigation and Methodology: PKA, IM, SMS, TC; Project administration: TC, PKA, SMS; Resources, software, validation, visualisation: PKA, SMS; Writing – review & editing: TC, PKA, SMS, GO.

## Ethical Considerations

This study did not require new approval from the institutional ethics committee, as it did not involve any clinical interventions, and participation was entirely voluntary and anonymous. Human participants were involved in both expert evaluations and user studies. All participants participated voluntarily, and informed consent was obtained prior to participation. The user study was conducted using an interactive chatbot interface in a controlled research setting, where participants were shown a consent notice outlining the study’s purpose, data usage, and privacy safeguards. No personally identifiable or sensitive information was collected at any stage.

## Data Sharing

The data supporting the findings of this study are publicly available at Hugging Face. The dataset can be accessed at https://huggingface.co/datasets/proadhikary/MENST. Additionally, the pre-trained models and code used in this study are hosted on Hugging Face Model Hub and can be accessed at https://huggingface.co/proadhikary/Menstrual-LLaMA-8B. For further details or to report any issues, please contact the corresponding author.

## Acknowledgements

TC would like to thank the financial support of the Tower Research Capital Markets toward using machine learning for social good, Rajiv Khemani Young Faculty Chair Professorship in AI, and the equipment support of central HPC facility (Padum).

## Conflict of interest

The authors have no conflict of interest.

## Abbreviations

AI: Artificial Intelligence
BERTScore: Bidirectional Encoder Representations from Transformers Score
BLEU: Bilingual Evaluation Understudy
GPT: Generative Pre-trained Transformer
JMIR: Journal of Medical Internet Research
LLM: Large Language Model
METEOR: Metric for Evaluation of Translation with Explicit Ordering
MHE: Menstrual Health Education
NLP: Natural Language Processing
PEFT: Parameter Efficient Fine-Tuning
QA: Question Answering
ROUGE-L: Recall-Oriented Understudy for Gisting Evaluation – Longest Common Subsequence

## Notes

### Competing Interest Statement

The authors have declared no competing interest.

### Funding Statement

We would like to thank the financial support of the Tower Research Capital Markets toward using machine learning for social good, Rajiv Khemani Young Faculty Chair Professorship in AI, and the equipment support of central HPC facility (Padum).

### Summary of Updates

The abstract was expanded to 450 words with added numerical results and formatting fixes (including metadata update); the introduction was revised to critique prior chatbot systems, highlight MenstLLaMAs educational focus, and clearly state the study aim; methods were updated with details on expert validation, inter-rater reliability, and participant recruitment and compensation; demographic data and evaluator sourcing were added; Figure 2 was revised for clarity; all figures were enhanced; the discussion now includes deeper insights and proposed next steps; limitations were expanded to address biases, misinformation, ethical risks, and privacy concerns; data security practices were clarified; evaluation metrics were contextualized for the health domain; tables were reformatted per JMIR guidelines; AI usage disclosure was included; and a tracked-changes version was uploaded as a supplementary file.

